# Effectiveness of high-dose versus standard-dose influenza vaccines against severe respiratory and cardiovascular outcomes in adults aged ≥80 years, Andalusia, Spain, 2024-2025 season

**DOI:** 10.64898/2025.12.20.25342573

**Authors:** Laura Díaz-Estévez, Mario Rivera-Izquierdo, Nicolás Francisco Fernández-Martínez, Daniel Ocaña-Rodríguez, David Moreno-Pérez, Nicola Lorusso

## Abstract

**Background:** Vaccination is a key measure to prevent severe influenza in adults aged ≥80 years, who experience the highest burden of respiratory and cardiovascular complications. Because immunosenescence reduces the effectiveness of standard-dose vaccines (SD-IIV), enhanced formulations such as high-dose vaccines (HD-IIV) are recommended in the elderly. Real-world data on their effectiveness in adults aged ≥80 years remain scarce.

**Aim:** To quantify the relative vaccine effectiveness (rVE) of HD-IIV versus SD-IIV in preventing severe influenza disease in adults aged ≥80 years.

**Methods:** Retrospective, population-based cohort study conducted in Andalusia, Spain, during the 2024-2025 influenza season, including 279,649 vaccinated adults aged ≥80 years. Data on sociodemographic characteristics, influenza vaccines, chronic diseases and clinical outcomes were taken from the Andalusian health population database. We used a directed acyclic graph to illustrate the assumed relationships between variables. The rVE of HD-IIV versus SD-IIV was estimated using augmented inverse probability weighting models.

**Results:** Compared with SD-IIV, HD-IIV was associated with a lower risk of hospitalization for influenza (rVE=34.0%; 95% CI=15.8-52.2). HD-IIV also showed improved effectiveness against laboratory-confirmed influenza (rVE=43.1%; 95% CI=24.6-61.7), hospitalization for acute myocardial infarction (rVE=26.4%; 95% CI=5.6-47.2), for stroke (rVE=32.9%; 95% CI=17.4-47.4), for pulmonary embolism (rVE=26.7%; 95% CI=0.7-52.6) and for overall cardiovascular outcomes (rVE=6.6%; 95% CI=0.9-12.2). No association was observed for hospitalization due to pneumonia, influenza/pneumonia, heart failure, respiratory outcomes, or in-hospital mortality.

**Conclusion:** Among adults aged ≥80 years, HD-IIV was more effective than SD-IIV in preventing hospitalization for influenza and severe cardiovascular outcomes, supporting its use in this at-risk population.

## Background

Influenza remains a leading vaccine-preventable cause of morbidity and mortality, as it accounts for 290,000–650,000 annual deaths globally [1]. An estimated 15,000–70,000 deaths per year are attributable to influenza in European countries, with a disproportionately high mortality burden in older age groups [2]. Epidemiologic surveillance data from Spain indicate that, in the last influenza seasons, population aged 80 and above show the highest rates of influenza-associated pneumonia, hospitalization, and mortality [3]. Besides, its clinical impact extends beyond respiratory illness–critically, there is growing recognition of influenza as a trigger for acute cardiovascular events in older adults, including myocardial infarction, heart failure and stroke [4].

Consequently, influenza vaccination constitutes a priority public health intervention and is systematically recommended for at risk groups, including older adults [5]. However, the most commonly employed standard-dose vaccine, with 15 µg of hemagglutinin per virus strain [6], has raised concerns owing to its limited effectiveness in certain populations. Both the magnitude and duration of the humoral response to influenza vaccination are reduced in the elderly [7]. High-dose influenza vaccines containing 60 µg of hemagglutinin have been developed to induce a more robust and sustained response in adults aged ≥60 years [8]. Compared to standard-dose, high-dose influenza vaccines have shown a relative vaccine effectiveness (rVE) between 5.7% and 8.4% in preventing cardiorespiratory hospitalization in adults aged ≥65 years [9–11]. This protection encompasses the prevention of viral infection and the attenuation of downstream cardiovascular and respiratory events. However, evidence from adults aged 80 years and above is scarce, as this age group often underrepresented (if not excluded).

Andalusia is the most populated region in Spain (with more than 8.5 million inhabitants as of 2024). The 2024-2025 influenza season in Andalusia elapsed from October to March and, in early January, it reached the maximum incidence rate (estimated at 276 cases per 100,000 population), substantially higher than in the previous two seasons [12]. This season was dominated by influenza A, with H3N2 being the predominant circulating subtype [12]. The influenza vaccination campaign in the 2024-2025 season included the following vaccines in adults aged ≥80 years: i) high-dose, tetravalent, egg-culture, inactivated vaccine; ii) standard-dose, tetravalent, egg-culture, inactivated vaccine; and c) standard-dose, tetravalent, cell-culture, inactivated vaccine [13]. This campaign was largely based on results from high-quality randomized clinical trials. Yet, influenza vaccination campaigns could benefit from real-world evidence: there is a need to evaluate the effect of administering the high-dose vaccine to adults aged 80 years and older on cardiovascular and respiratory outcomes.

Therefore, to fill in this knowledge gap, the objective of this study was to quantify the rVE of high-dose vs. standard-dose influenza vaccination in preventing severe influenza in individuals aged 80 years and above.

## Methods

### Study design and population

We conducted an observational, retrospective, population-based cohort study. The study period ranged from 9 October, 2024 (week 41)–the start date of the general influenza vaccination campaign in Andalusia–to 16 March, 2025 (week 11), 28 days after the end of the regional epidemic, according to national epidemic thresholds calculated using the moving epidemic method (MEM) [14]. We chose this follow-up period because it is a commonly used time frame to evaluate early complications plausibly related to influenza (longer time intervals may capture additional events at the expense of specificity). The study population consisted of adults aged ≥80 years as of 9 October, 2024, listed as users of the Andalusian Public Healthcare System (a tax-based system with virtually universal coverage). Participants in whom the effect of influenza vaccination could not be assessed (i.e., diagnosed with influenza prior to vaccination or within 14 days following vaccination), participants who received more than one influenza vaccine, and participants who received an unknown type of influenza vaccine were excluded.

### Data sources

Data were obtained from an anonymized dataset provided by the Andalusian health population database [15], which comprises information from multiple health records, including, but not limited to, sociodemographic characteristics, vaccines, laboratory and diagnostic imaging tests, chronic diseases, hospital admissions and healthcare utilization (it should be noted that this database does not include information from private healthcare centers) [16]. Diagnostic data were coded using the International Classification of Diseases 9^th^ (ICD-9) and 10^th^ edition (ICD-10).

### Variables and definitions

Variables included in this study are detailed in Supplementary Table S1. The primary exposure (main independent variable) was the type of influenza vaccination received: high-dose, tetravalent, cell-culture, inactivated influenza vaccine (HD-IIV) (Efluelda Tetra®, Sanofi) vs. standard-dose, tetravalent, either cell- or egg-culture, inactivated influenza vaccine (SD-IIV) (Flucelvax Tetra®, Seqirus or Vaxigrip Tetra®, Sanofi, respectively). The virus-like strains contained in these vaccines are shown in Supplementary Table S2.

Study outcomes (dependent variables) were chosen following the protocol of the DANFLU-1 trial [17] where possible, in an attempt to improve comparability. Our primary outcomes were related to severe influenza disease and comprised:

– Hospitalization for influenza.
– Hospitalization for pneumonia.
– Hospitalization for influenza/pneumonia (both combined).

As secondary outcomes, we selected:

– Laboratory-confirmed influenza (detected by polymerase chain reaction).
– Hospitalization for acute myocardial infarction.
– Hospitalization for stroke.
– Hospitalization for pulmonary embolism.
– Hospitalization for heart failure.
– Hospitalization for respiratory outcomes.
– Hospitalization for cardiovascular outcomes.
– All-cause in-hospital mortality.

In addition, hospitalization for osteoporotic fractures/hip replacement surgery was included as a negative control outcome. We assumed that this outcome was unassociated with the exposure (thereby expecting to yield a null rVE in the absence of bias), although we acknowledge the theoretical small association between influenza-induced falls and fractures.

Vaccine effectiveness (VE) was defined the reduction in the risk of developing each of the study outcomes, calculated as VE (%) = (1 - risk ratio) x 100.

Following expert recommendations [18], we used a directed acyclic graph (DAG) to guide the selection of covariates (Figure 1) and we stated our estimands (specified below). The covariates included sociodemographic factors (age, sex, province, health district, area-level socioeconomic status), institutionalization, healthcare utilization, date of vaccination, and baseline chronic diseases. To comprehensively capture chronic diseases in the study population, we used ICD-9 and ICD-10 codes with a 35-year look-back period.

**Figure 1.**
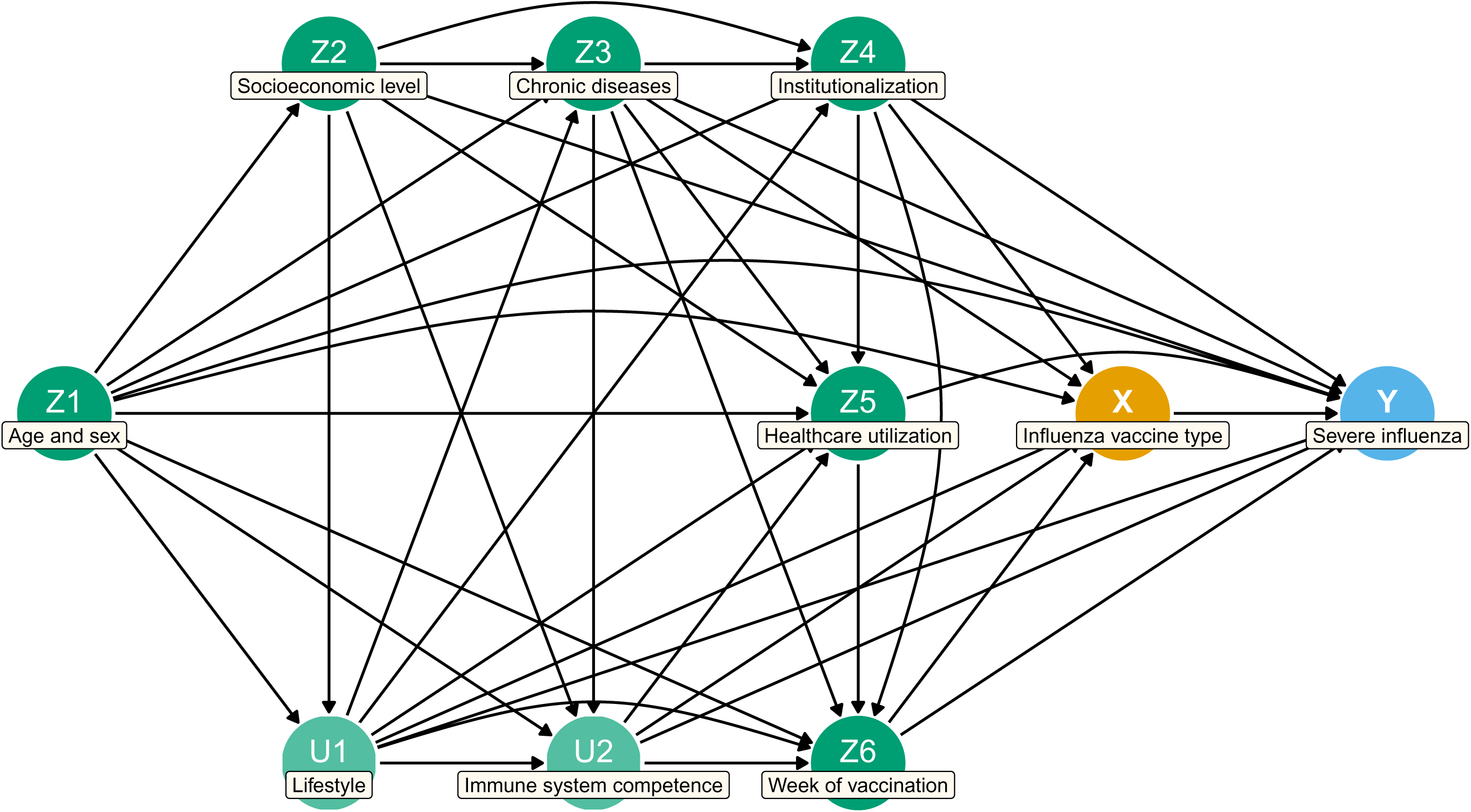
Directed acyclic graph illustrating the assumptions about the causal relationships between variables.

### Statistical analysis

We described the characteristics of the study participants using absolute and relative (%) frequencies for qualitative variables and if median and interquartile range (IQR) for quantitative variables (since they did not follow the normal distribution). In the bivariate analysis, we compared participants by type of vaccination using the Chi-square test and the Kruskal-Wallis test for qualitative and quantitative variables, respectively.

Our estimands were the rVE of influenza vaccination in preventing each of the study outcomes in participants who received HD-IIV compared to those who received SD-IIV. To quantify the degree of bias in our estimates, our estimand was the rVE of influenza vaccination in preventing hospitalization for osteoporotic fractures/hip replacement surgery in participants who received HD-IIV compared to those who received SD-IIV.

Doubly-robust augmented inverse probability weighting (aIPW) models were built to obtain adjusted risk ratios alongside their 95% confidence intervals (CI). All models used to obtain rVE estimates were adjusted for age, sex, location of the health center in a socioeconomically deprived area, institutionalization, baseline chronic diseases (heart failure, dementia, neurological/neuromuscular disorders and cerebrovascular disease), number of visits to primary care in the previous five years, and week of vaccination. As a sensitivity analysis, we employed multivariable log-binomial regression models (Supplementary Table S3). Continuous covariates were not categorized. We verified that the conditions of application of statistical models were met. All tests were two-sided and type 1 (α) error was set at 5%. All analyses were performed using R v4.4.1 (R Core Team, https://www.R-project.org) and Stata v15.1 (Stata Statistical Software: Release 15. College Station, TX: StataCorp LP). Figures were produced using the ggplot2 and ggdag R packages.

## Results

### Description of the study sample and the vaccination campaign

The study sample comprised 279,649 participants, including 207,421 vaccinated with HD-IIV and 72,228 vaccinated with SD-IIV. About two thirds of SD-IIV recipients were administered egg-culture vaccines (48,890, 67.7%). Median age of participants was 85.0 years (IQR=82.2–88.8 years) and 61.3% were women. The frequency of high-dose vaccination increased with age (from 70.4% in the 80–84 age group to 88.2% in those aged ≥100 years). Institutionalization was also more common in participants who received HD-IIV (7.2% vs. 0.9%). Compared to SD-IIV, individuals vaccinated with HD-IIV had a higher median of primary care visits in the previous years (97 vs. 88). Participants receiving HD-IIV had a higher frequency of certain chronic diseases at baseline, namely heart failure (20.8% vs. 19.0%), dementia (15.0% vs. 10.2%), neurological/neuromuscular disorders (8.2% vs. 7.5%) and cerebrovascular disease (6.1% vs. 5.6%). The baseline characteristics of study participants by exposure group are presented in Table 1.

**Table 1.** Baseline characteristics of the participants vaccinated with high-dose vs. standard-dose influenza vaccines, Andalusia, Spain, 2024-2025 season (n = 279,649).

| Characteristic | HD-IIV<br>n = 207,421 (74.2%) |  | SD-IIV<br>n = 72,228 (25.8%) |  | p value |
| --- | --- | --- | --- | --- | --- |
|  | Number | Row % | Number | Row % |  |
| *Sex* |  |  |  |  | <0.001<sup>1</sup> |
| Male | 78,675 | 72.7% | 29,511 | 27.3% |  |
| Female | 128,746 | 75.1% | 42,717 | 24.9% |  |
| *Age group (years)* |  |  |  |  | <0.001<sup>1</sup> |
| 80–84 | 98,701 | 70.4% | 41,518 | 29.6% |  |
| 85–89 | 65,970 | 76.1% | 20,668 | 23.9% |  |
| 90–94 | 34,425 | 80.3% | 8,458 | 19.7% |  |
| 95–99 | 7,503 | 83.6% | 1,474 | 16.4% |  |
| ≥100 | 822 | 88.2% | 110 | 11.8% |  |
| *Province* |  |  |  |  | <0.001<sup>1</sup> |
| Almería | 15,131 | 76.7% | 4,608 | 23.3% |  |
| Cádiz | 23,328 | 70.0% | 10,006 | 30.0% |  |
| Córdoba | 28,173 | 80.5% | 6,806 | 19.5% |  |
| Granada | 24,442 | 67.8% | 11,598 | 32.2% |  |
| Huelva | 10,168 | 63.2% | 5,926 | 36.8% |  |
| Jaén | 22,414 | 78.2% | 6,264 | 21.8% |  |
| Málaga | 36,638 | 75.8% | 11,708 | 24.2% |  |
| Sevilla | 47,127 | 75.5% | 15,312 | 24.5% |  |
| *Institutionalization* | 15,017 | 96.0% | 618 | 4.0% | <0.001<sup>1</sup> |
| *Socioeconomically deprived area* | 42,119 | 74.5% | 14,443 | 25.5% | 0.075<sup>1</sup> |
| *Primary care visits in the previous 5 years (median, IQR)* | 97 | 65–144 | 88 | 59–129 | <0.001<sup>2</sup> |
| *Chronic cardiovascular disease* | 184,075 | 74.3% | 63,793 | 25.7% | 0.002<sup>1</sup> |
| *Arterial hypertension* | 176,781 | 74.3% | 61,281 | 25.7% | 0.013<sup>1</sup> |
| *Heart failure* | 43,232 | 75.9% | 13,692 | 24.1% | <0.001<sup>1</sup> |
| *Atrial fibrillation* | 39,220 | 75.2% | 12,964 | 24.8% | <0.001<sup>1</sup> |
| *Valvular heart disease* | 27,934 | 74.6% | 9,531 | 25.4% | 0.066<sup>1</sup> |
| *Ischemic heart disease* | 27,584 | 74.1% | 9,645 | 25.9% | 0.713<sup>1</sup> |
| *Cerebrovascular disease* | 12,664 | 75.7% | 4,068 | 24.3% | <0.001<sup>1</sup> |
| *Chronic lung disease* | 49,722 | 74.4% | 17,111 | 25.6% | 0.128<sup>1</sup> |
| *Chronic obstructive pulmonary disease* | 34,391 | 74.4% | 11,830 | 25.6% | 0.211<sup>1</sup> |
| *Asthma* | 22,755 | 74.3% | 7,870 | 25.7% | 0.586<sup>1</sup> |
| *Diabetes* | 79,499 | 74.5% | 27,194 | 25.5% | 0.001<sup>1</sup> |
| *Dementia* | 31,028 | 80.9% | 7,339 | 19.1% | <0.001<sup>1</sup> |
| *Neurological/neuromuscular disorder* | 17,100 | 75.9% | 5,419 | 24.1% | <0.001<sup>1</sup> |
| *Chronic kidney disease* | 25,902 | 75.1% | 8,607 | 24.9% | <0.001<sup>1</sup> |
| *Rheumatic disease* | 10,711 | 75.0% | 3,567 | 25.0% | 0.018<sup>1</sup> |
| *Chronic liver disease* | 8,698 | 73.5% | 3,128 | 26.5% | 0.117<sup>1</sup> |
| *Prostate cancer* | 8,867 | 72.6% | 3,347 | 27.4% | <0.001<sup>1</sup> |
| *Colorectal cancer* | 7,031 | 74.3% | 2,435 | 25.7% | 0.823<sup>1</sup> |
| *Breast cancer* | 6,226 | 74.3% | 2,149 | 25.7% | 0.730<sup>1</sup> |
| *Obesity* | 29,587 | 73.9% | 10,472 | 26.1% | 0.123<sup>1</sup> |
| *Date of vaccination (week of campaign)* |  |  |  |  | <0.001<sup>1</sup> |
| Weeks 1 and 2 | 123,755 | 87.3% | 18,024 | 12.7% |  |
| Weeks 3 and 4 | 47,995 | 70.6% | 19,961 | 29.4% |  |
| Weeks 5 and 6 | 21,503 | 56.9% | 16,288 | 43.1% |  |
| Weeks $\geq 7$ | 14,168 | 44.1% | 17,955 | 55.9% | |

Data are presented as absolute frequency (n) and relative frequency (%) for qualitative variables, and as median and interquartile range (IQR) for quantitative variables. HD-IIV: high-dose, inactivated influenza vaccine. SD-IIV: standard-dose, inactivated influenza vaccines. <sup>1</sup>p values from chi-square test. <sup>2</sup>p values from Mann-Whitney U test.

The influenza vaccination campaign commenced on 9 October, 2024. Differentiated phases applied for each group at risk (e.g., vaccination of individuals aged 60 to 69 years did not start until 23 October), although the start date of vaccination was the same for all adults ≥80 years (9 October). Vaccine administration was concentrated in the early stages of the campaign, with 141,779 participants (50.7%) vaccinated in the first two weeks. However, the distribution of vaccine types was not uniform throughout the study period. While HD-IIV was the main vaccine at the start of the campaign, its relative frequency declined significantly over time: from the seventh week onwards, it was surpassed by SD-IIV. The timeline of the vaccination campaign is represented in Figure 2.

**Figure 2.**
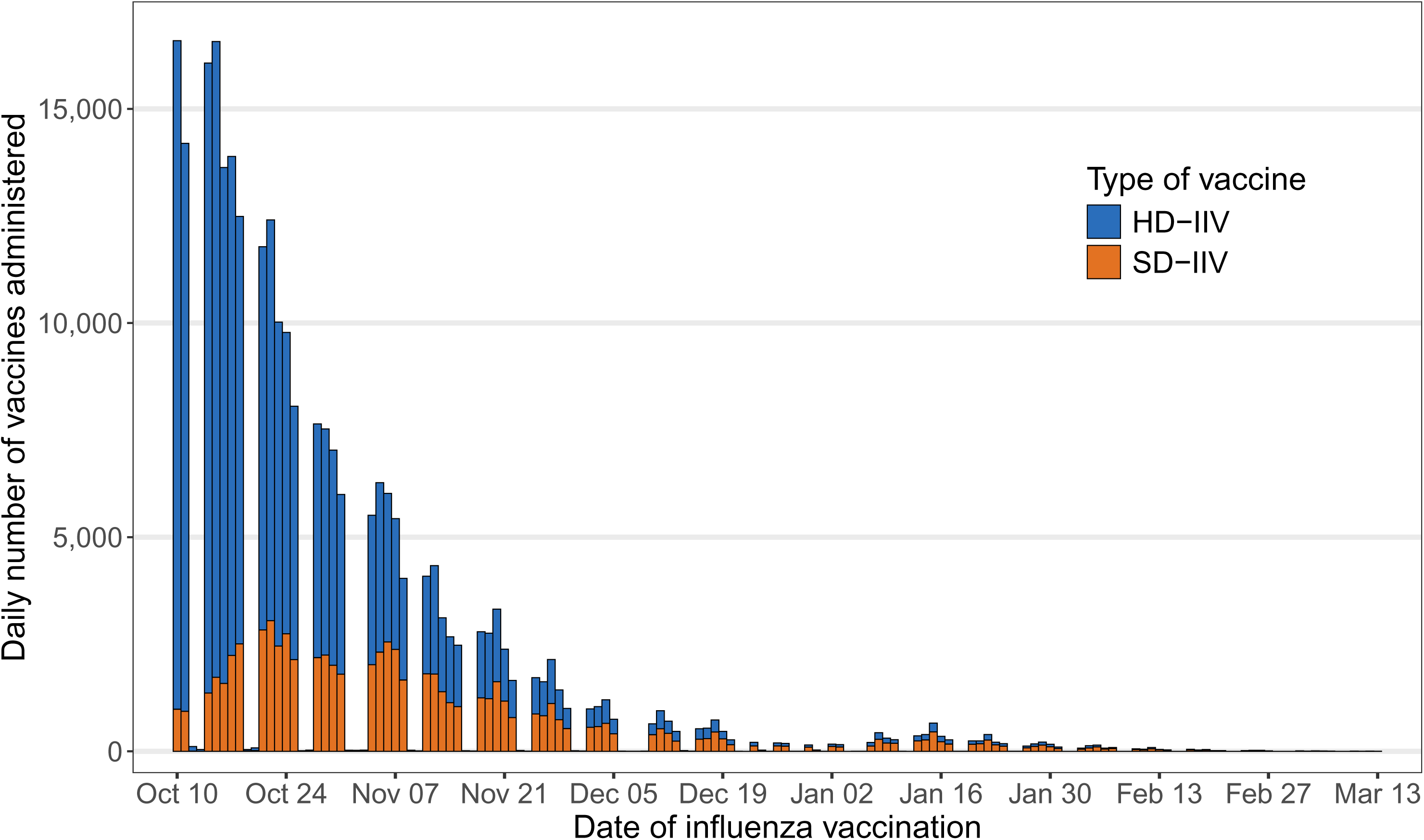
Histogram displaying the timeline of the 2024-2025 influenza vaccination campaign in Andalusia, Spain, by vaccine type (n = 279,649). Figure note: HD-IIV: high-dose, inactivated influenza vaccine. SD-IIV: standard-dose, inactivated influenza vaccines.

### Description of study outcomes

Table 2 shows the distribution of primary and secondary outcomes by exposure group. A total of 642 participants (0.2%) were hospitalized for influenza, 2,117 (0.8%) for pneumonia, and 2,534 (0.9%) for influenza/pneumonia. Adults who were administered HD-IIV had a higher occurrence of hospitalization for pneumonia (0.8% vs. 0.6%), for respiratory (2.6% vs. 2.0%) and for cardiovascular outcomes (3.8% vs. 3.2%), as well as a higher in-hospital mortality (0.3% vs. 0.2%). In contrast, the incidence of laboratory-confirmed influenza–mostly, influenza A, which accounted for 95.1% of all cases–was lower in participants in the HD-IIV group.

**Table 2.** Incidence of study outcomes in the participants vaccinated with high-dose vs. standard-dose influenza vaccines, Andalusia, Spain, 2024-2025 season (n = 279,649).

| Outcome | HD-IIV<br>n = 207,421<br>(74.2%) |  | SD-IIV<br>n = 72,228 (25.8%) |  | p<br>value |
| --- | --- | --- | --- | --- | --- |
|  | Number | Column % | Number | Column % |  |
| Primary outcomes (severe influenza) |  |  |  |  |  |
| Hospitalization for influenza | 482 | 0.2% | 160 | 0.2% | 0.631<sup>1</sup> |
| Hospitalization for pneumonia | 1,696 | 0.8% | 421 | 0.6% | <0.001<sup>1</sup> |
| Hospitalization for influenza/pneumonia | 2,006 | 1.0% | 528 | 0.7% | <0.001<sup>1</sup> |
| Secondary outcomes |  |  |  |  |  |
| Laboratory-confirmed influenza | 265 | 0.1% | 119 | 0.2% | 0.024<sup>1</sup> |
| Influenza A | 253 | 0.1% | 112 | 0.2% | 0.039<sup>1</sup> |
| Influenza B | 12 | <0.1% | 7 | <0.1% | 0.295<sup>2</sup> |
| Hospitalization for acute myocardial infarction | 309 | 0.1% | 128 | 0.2% | 0.110<sup>1</sup> |
| Hospitalization for stroke | 461 | 0.2% | 185 | 0.3% | 0.112<sup>1</sup> |
| Hospitalization for pulmonary embolism | 194 | 0.1% | 73 | 0.1% | 0.621<sup>1</sup> |
| Hospitalization for heart failure | 1,483 | 0.7% | 481 | 0.7% | 0.183<sup>1</sup> |
| Hospitalization for respiratory outcomes | 5,364 | 2.6% | 1,470 | 2.0% | <0.001<sup>1</sup> |
| Hospitalization for cardiovascular outcomes | 7,782 | 3.8% | 2,312 | 3.2% | <0.001<sup>1</sup> |
| In-hospital mortality | 545 | 0.3% | 138 | 0.2% | <0.001<sup>1</sup> |

Data are presented as absolute frequency (n) and relative frequency (%). HD-IIV: high-dose, inactivated influenza vaccine. SD-IIV: standard-dose, inactivated influenza vaccines. <sup>1</sup>p values from chi-square test. <sup>2</sup>p values from Fisher's exact test.

### Relative vaccine effectiveness (rVE)

HD-IIV was associated with a reduction in the risk of hospitalization for influenza (rVE=34.0%, 95% CI=15.8% to 52.2%). No association was found between vaccination with HD-IIV vs. SD-IIV and hospitalization for pneumonia, or hospitalization for influenza/pneumonia. Regarding pre-specified secondary outcomes, HD-IIV was associated with a lower risk of laboratory-confirmed influenza (rVE=43.1%; 95% CI=24.6% to 61.7%). High-dose vaccination was further associated with a decreased risk of hospitalization for the following cardiovascular outcomes: acute myocardial infarction (rVE=26.4%; 95% CI=5.6% to 47.2%), stroke (rVE=32.9%; 95% CI=17.4% to 47.4%), pulmonary embolism (rVE=26.7%; 95% CI=0.7% to 52.6%), and overall cardiovascular outcomes (rVE=6.6%, 95% CI=0.9% to 12.2%). Receiving HD-IIV was not associated with hospitalization for heart failure, respiratory outcomes, or in-hospital mortality.

No association was observed between HD-IIV and hospitalization for osteoporotic fractures/hip replacement surgery in the negative control analysis. The crude rVE of HD-IIV vs. SD-IIV was - 45.4% (95% CI=-73.9% to -22.3%), which increased to an adjusted rVE of -6.4% (95% CI=-32.6% to 19.8%) after multivariable adjustment. This finding indicates that the values herein obtained were not found to be significantly biased. The rVE of HD-IIV vs. SD-IIV against study outcomes is displayed in Figure 3 and detailed in Table 3.

**Figure 3.**
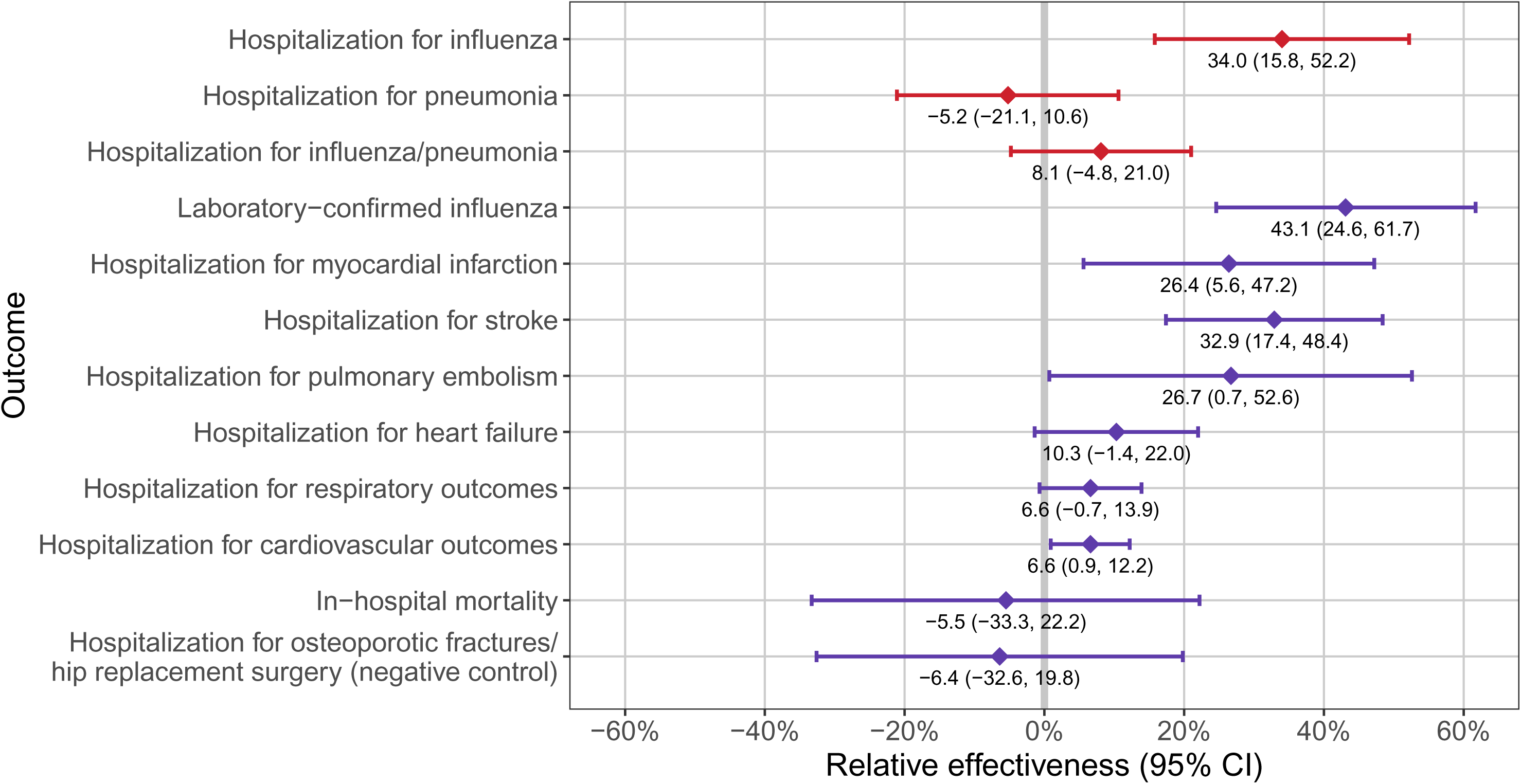
Relative vaccine effectiveness estimates of HD-IIV vs. SD-IIV in preventing primary (red) and secondary (purple) study outcomes, Andalusia, Spain, 2024-2025 season (n = 279,649). Figure note: HD-IIV: high-dose, inactivated influenza vaccine. SD-IIV: standard-dose, inactivated influenza vaccines.

**Table 3.** Relative vaccine effectiveness of high-dose (HD-IIV) vs. standard-dose (SD-IIV) influenza vaccines on study outcomes, Andalusia, Spain, 2024-2025 season (n = 279,649).

| Outcome | rVE (%) | 95% CI (%) |
| --- | --- | --- |
| **Primary outcomes (severe influenza)** |  |  |
| *Hospitalization for influenza* | 34.0 | (15.8 to 52.2) |
| *Hospitalization for pneumonia* | -5.2 | (-21.1 to 10.6) |
| *Hospitalization for influenza/pneumonia* | 8.1 | (-4.8 to 21.0) |
| **Secondary outcomes** |  |  |
| *Laboratory-confirmed influenza* | 43.1 | (24.6 to 61.7) |
| Influenza A | 43.3 | (24.2 to 62.4) |
| Influenza B | 38.8 | (-24.8 to 102.5) |
| *Hospitalization for acute myocardial infarction* | 26.4 | (5.6 to 47.2) |
| *Hospitalization for stroke* | 32.9 | (17.4 to 48.4) |
| *Hospitalization for pulmonary embolism* | 26.7 | (0.7 to 52.6) |
| *Hospitalization for heart failure* | 10.3 | (-1.4 to 22.0) |
| *Hospitalization for respiratory outcomes* | 6.6 | (-0.7 to 13.9) |
| *Hospitalization for cardiovascular outcomes* | 6.6 | (0.9 to 12.2) |
| *In-hospital mortality* | -5.5 | (-33.3 to 22.2) |
| *Hospitalization for osteoporotic fractures/hip replacement surgery (negative control outcome)* | -6.4 | (-32.6 to 19.8) |

Doubly-robust augmented inverse probability weighting (aIPW) models adjusted for age, sex, location of the health center in an area in needs of social transformation, institutionalization, baseline chronic diseases (heart failure, dementia, neurological/neuromuscular disorders and cerebrovascular disease), number of visits to primary care in the previous five years, and week of vaccination. CI: confidence interval. HD-IIV: high-dose, inactivated influenza vaccine. rVE: relative vaccine effectiveness. SD-IIV: standard-dose, inactivated influenza vaccines.

## Discussion

In this retrospective, population-based cohort study of adults aged 80 years and above, vaccination with HD-IIV vs. SD-IIV appeared to reduce hospitalization for influenza (rVE=34%). Moreover, HD-IIV was associated with a lower incidence of secondary outcomes related to influenza risk and hospitalization for cardiovascular diseases, including laboratory-confirmed influenza (rVE=43%), hospitalization for acute myocardial infarction (rVE=26%), for stroke (rVE=33%), pulmonary embolism (rVE=27%) and cardiovascular outcomes (rVE=7%). Compared to SD-IIV, HD-IIV did not have a higher rVE on hospitalization for pneumonia, hospitalization for influenza/pneumonia, hospitalization for respiratory outcomes, or all-cause in-hospital mortality.

Our main finding is aligned with that from previous studies (with the caveat that reports from previous research usually apply to slightly younger age groups). In the pragmatic, open-label, randomized, controlled DANFLU-2 trial conducted in Denmark between the 2022-2023 and the 2024-2025 influenza seasons, HD-IIV vs. SD-IIV showed a protective effect against hospitalization for influenza (rVE=44%) [19]. However, no association between HD-IIV and hospitalization for pneumonia was found [19]. Later, Martinón-Torres et al. conducted GALFLU, a pragmatic, open-label, randomized, controlled trial in Galicia, Spain, covering the 2023-2024 and 2024-2025 seasons. They found that HD-IIV decreased the risk of hospitalization for influenza by 32% compared to SD-IIV, whereas the effect on hospitalization for pneumonia was non-significant [10]. In fact, in the recently published FLUNITY-HD trial, a pooled analysis of DANFLU-2 and GALFU, a lower incidence of hospitalization for influenza, but not hospitalization for pneumonia, was observed for HD-IIV (rVE=40% and 2%, respectively) [11].

Moreover, similar findings have also been reported in observational studies. A retrospective, community-based cohort study conducted in France during the 2021-2022 season observed an association between vaccination with HD-IIV vs. SD-IIV and hospitalization for influenza (rVE=23%), but not hospitalization for pneumonia (rVE=0%) [20]. A retrospective cohort study conducted in the United States that analyzed six consecutive influenza seasons concluded that HD-IIV was associated with a slight decrease in the risk of hospitalization for influenza (pooled rVE=14%) and, to a lesser extent, hospitalization for pneumonia (pooled rVE=4%) [21].

Although the observed rVE of vaccination with HD-IIV was relatively modest, it represents the added benefit attributable to high-dose vaccines on top of standard-dose vaccines. Notably, this effect was demonstrated in a susceptible population characterized by weaker immune response to influenza vaccines, which could bias comparisons of HD-IIV vs. SD-IIV toward the null. Thus, even a small effect size may hold meaningful clinical relevance in this context.

On the other hand, vaccination with HD-IIV offered protection against several secondary outcomes. The highest effect (rVE=43%) was observed for laboratory-confirmed influenza, in line with previous research [22,23]. In light of the historical challenges posed by the influenza vaccine mismatch with H3N2 (the predominant A subtype during the 2024-2025 season in Andalusia), this finding underscores the ability of HD-IIV to broaden the immune repertoire, even against drift variants, and indicates that a higher antigen dose could correctly address the limitations of SD-IIV against H3N2. Furthermore, we found that HD-IIV seemed to lower the risk of hospitalization for cardiovascular outcomes, including acute myocardial infarction, stroke and pulmonary embolism (of note, the 95% CI lower limit of the latter was estimated at <1%, very close to the null). This suggests that the previously established link between HD-IIV and a reduced risk of cardiovascular hospitalization in older adults [24,25] is also valid for the population aged ≥80 years. However, to our knowledge, protection against stroke (rVE=33%) is a novel finding from our study with potential implications for clinical practice. Influenza exerts direct vascular effects and a systemic inflammatory response, which can lead to hypoxemia, atherosclerotic plaque accumulation and instability, and hypercoagulability [26]. Enhanced influenza vaccines may mitigate the intensity of this response and reduce the occurrence of stroke. This finding adds up to the body of evidence supporting the preventive effect of HD-IIV on cardiovascular outcomes.

With the above considerations, the fact that HD-IIV protected against specific outcomes (vs. SD-IIV) could mean that older adults benefit from the high-dose vaccines particularly in terms of prevention of influenza and its direct complications. From a biological standpoint, this finding can be explained by immunosenescence, a phenomenon of immune dysfunction associated with aging [27]. The rationale for the development of high-dose influenza vaccines for older adults was precisely to compensate the lower immunogenicity of standard-dose vaccines. Accordingly, it is not unexpected to find higher differences between both vaccine types regarding outcomes strictly attributable to influenza. Due to their higher specificity, these are regarded as more appropriate to assess the effectiveness of influenza vaccines [28]. For broader outcomes, admittedly more relevant to public health, the effect of HD-IIV may be diminished, because non-specific endpoints are more influenced by individual (e.g., all-cause in-hospital mortality depends on comorbidity and frailty [29]) or epidemiological factors (e.g., protection against hospitalization for pneumonia depends on the proportion of pneumonia cases attributable to influenza and relies on the accuracy of diagnostic codes for composite outcomes in this population), among others. However, this did not apply to cardiovascular outcomes in this study, with which HD-IIV was inversely associated. It is also plausible that we may have not been able to replicate the protective effect of HD-IIV on certain outcomes due to inherent flaws with the observational design.

The strengths of this study include a cohort design with access to population-based data, which allowed us to analyze multiple outcomes (including cardiovascular events), a large sample size, and the application of a causal framework through DAGs and inverse probability weighting models. This study also has some limitations. First, the follow-up period, which was until mid-March, is shorter in length than that of comparable studies, resulting in a smaller number of events observed (and thus a lower statistical power). Nevertheless, in doing so, we estimated the effectiveness of vaccination with HD-IIV vs. SD-IIV during the influenza epidemic, a critical period due to the high burden on the healthcare system. Second, because of limited statistical power, we could not assess in-hospital mortality due to influenza or its complications. We analyzed all-cause mortality instead, which can be influenced by myriad causes other than influenza. Third, a certain degree of underdetection of study outcomes cannot be ruled out, given the retrospective design and the use of a secondary data source. This was likely to happen in both exposure groups, producing a non-differential bias that probably resulted in an underestimation of the rVE of HD-IIV. Fourth, the SD-IIV group included two different vaccines (egg-culture and cell-culture). While this decision introduced heterogeneity, we considered them jointly as a single group because, in the study region, both are used interchangeably as standard influenza vaccines with the same indications. Fifth, our results are likely subject to residual confounding. Despite our best efforts to address this, we were not able to control the effect of some confounding factors (e.g., immunosuppression). Multivariable analysis substantially reduced this bias, which was non-significant after adjustment, but still indicated that we underestimated the rVE of HD-IIV in about six percentage points. Sixth, influenza vaccination has been associated to lower risk of ischemic, but not hemorrhagic stroke, whereas we analyzed stroke as a whole. Future studies assessing this outcome should report effectiveness estimates stratified by type of stroke. Seventh, the inclusion of adults ≥80 years who were users of the public healthcare system only limited the generalizability of our findings. Yet, in this age group, non-users of the public system represent a small minority of the Andalusian population.

The practical implications of estimating the rVE of vaccination with HD-IIV vs. SD-IIV cannot be overstated–it is useful for the elderly (optimizing protection in a highly susceptible and understudied population), for health professionals and health policy evaluation (potentially improving the effectiveness of upcoming vaccination campaigns), and for economic evaluation (serving as a starting point for future cost-effectiveness analyses). To improve research in the observational context, future studies should incorporate a causal approach, assess additional enhanced influenza vaccine types (e.g., adjuvanted vaccines) and include outcomes beyond infection, hospitalization and death (such as quality of life, long-term sequelae and institutionalization).

In summary, this study found that, compared to its standard-dose counterparts, the high-dose influenza vaccine was effective in preventing hospitalization for influenza, laboratory-confirmed influenza, and hospitalization for cardiovascular outcomes (namely, acute myocardial infarction and stroke), although no association was found for hospitalization for pneumonia, hospitalization for influenza/pneumonia, hospitalization for respiratory outcomes, or in-hospital mortality. Our results on the added benefits of HD-IIV support its use in adults aged 80 years and above and showcase the importance of epidemiologic surveillance to inform vaccination campaigns.

## Data Availability

Data used in this study were obtained from the Andalusian health population database. The authors do not have permission to share the data.

