## Supplementary Tables 1-3 for "Effectiveness of high-dose versus standard-dose influenza vaccines against severe respiratory and cardiovascular outcomes in adults aged ≥80 years, Andalusia, Spain, 2024-2025 season"

Supplementary Table S1. Description of study variables.

| **Variable** | **Description** |
|---|---|
| **Independent variables** | |
| *Type of influenza vaccination* | High-dose, tetravalent, cell-culture, inactivated influenza vaccine (HD-IIV) or standard-dose, tetravalent, cell-/egg-culture, inactivated influenza vaccine (SD-IIV). |
| *Date of vaccination* | Date of vaccination with HD-IIV or SD-IIV. |
| *Age* | Age in years at the start of the study period. |
| *Sex* | Female or male biological sex. |
| *Province* | In Spain, the territory is organized into autonomous communities, which are divided into provinces. Andalusia encompasses eight provinces. |
| *Health district* | In Spain, the national health system is organized into health districts. Andalusia encompasses 34 health districts. |
| *Location of the health center in a socioeconomically deprived area* | In Andalusia, area-level socioeconomic deprivation is measured by "areas in need of social transformation". This classification by the Regional Government refers to urban areas defined by severe poverty and social exclusion. For more details, please see the Official Gazzette of the Andalusian Government no. 2 of 02/03/2016 (https://www.juntadeandalucia.es/boja/2016/502/). |
| *Institutionalization* | Institutionalization in a nursing home for elderly or disabled individuals. |
| *Baseline chronic diseases* | Chronic diseases or conditions present at baseline. Given the advanced age of study participants, a look-back period of 35 years was used.<br>The following diseases were included in the study (defined by ICD-9 and ICD-10 codes): chronic cardiovascular disease, arterial hypertension, heart failure, atrial fibrillation, valvular heart disease, ischemic heart disease, cerebrovascular disease and its sequelae, chronic lung disease, chronic obstructive pulmonary disease, asthma, diabetes, dementia, neurological/neuromuscular disorders, |

| | |
|---|---|
| | chronic kidney disease, rheumatic disease, chronic liver disease, prostate cancer, breast cancer (in females), colorectal cancer and obesity. |
| *Number of primary care visits in the previous five years* | Total number of visits to primary care in the five years prior to the 2024-2025 vaccination campaign. |
| **Dependent variables** | |
| Hospitalization for influenza | ICD-10 influenza codes J09-J11. |
| Hospitalization for pneumonia | ICD-10 pneumonia codes J09-J18. |
| Hospitalization for influenza/pneumonia | ICD-10 influenza and pneumonia codes J09-J18. |
| Laboratory-confirmed influenza | Influenza detected by PCR (polymerase chain reaction). |
| Hospitalization for acute myocardial infarction | ICD-10 acute myocardial infarction codes I21-I22. |
| Hospitalization for stroke | ICD-10 stroke codes I63-I64. |
| Hospitalization for pulmonary embolism | ICD-10 pulmonary embolism codes I26.0-I26.99. |
| Hospitalization for heart failure | ICD-10 heart failure codes I50.1-I50.43. |
| Hospitalization for respiratory outcomes | ICD-10 respiratory codes J00-J22, J40-J47, J80-J81, J85-J86, and J96. |
| Hospitalization for cardiovascular outcomes | ICD-10 cardiovascular codes I11, I13, I20-I25, I30, I31, I33, I38-I42, I46-I50, I60-I69, and I74. |
| In-hospital mortality | All-cause in-hospital mortality. |
| Hospitalization for osteoporotic fractures/hip replacement surgery | Negative control used to measure the degree of bias in estimates of vaccine effectiveness.<br>ICD-10 codes M80, M81.8, 0SR9, 0SRB, and those including osteoporosis or hip joint prothesis in the text description. |

Supplementary Table S2. Strains contained in the vaccines administered to adults aged ≥80 years in the 2024-2025 seasonal influenza campaign in Andalusia, Spain.

| **Vaccine** | **Strains** |
|---|---|
| High dose, egg-culture (Efluelda Tetra®)<br>Standard dose, egg-culture (Vaxigrip Tetra®) | A/Victoria/4897/2022 (H1N1)-like virus<br>A/Thailand /8/2022 (H3N2)-like virus<br>B/Austria/ 1359417/2021 (Victoria lineage)-like virus<br>B/Phuket/3073/2013 (Yamagata lineage)-like virus |
| Standard dose, cell-culture (Flucelvax Tetra®) | A/Wisconsin/67/2022 (H1N1)-like virus<br>A/Massachussetts/18/2022 (H3N2)-like virus<br>B/Austria/ 1359417/2021 (Victoria lineage)-like virus<br>B/Phuket/3073/2013 (Yamagata lineage)-like virus |

Supplementary Table S3. Relative vaccine effectiveness of high-dose (HD-IIV) vs. standard-dose (SD-IIV) influenza vaccines on study outcomes, Andalusia, Spain, 2024-2025 season. Sensitivity analysis.

| **Outcome** | **Regression model** | **rVE (%)** | **95% CI (%)** |
|---|---|---|---|
| **Primary outcomes (severe influenza)** | | | |
| *Hospitalization for influenza* | Log-binomial | 24.0 | (7.5 to 37.3) |
| *Hospitalization for pneumonia* | Log-binomial | -4.8 | (-17.6 to 6.4) |
| *Hospitalization for influenza/pneumonia* | Log-binomial | 2.2 | (-8.5 to 11.6) |
| **Secondary outcomes** | | | |
| *Laboratory-confirmed influenza* | Log-binomial | 39.3 | (22.8 to 52.0) |
| Influenza A | Log-binomial | 38.8 | (21.7 to 52.0) |
| Influenza B | Log-binomial | 47.1 | (-44.2 to 80.6) |
| *Hospitalization for acute myocardial infarction* | Log-binomial | 22.7 | (3.3 to 37.8) |
| *Hospitalization for stroke* | Log-binomial | 18.9 | (2.3 to 32.5) |
| *Hospitalization for pulmonary embolism* | Log-binomial | 22.1 | (-4.5 to 41.4) |
| *Hospitalization for heart failure* | Log-binomial | 7.8 | (-2.9 to 17.3) |
| *Hospitalization for respiratory outcomes* | Poisson* | 1.2 | (-5.1 to 7.1) |
| *Hospitalization for cardiovascular outcomes* | Poisson* | 2.2 | (-2.9 to 6.9) |
| *In-hospital mortality* | Log-binomial | 0.7 | (-21.5 to 18.4) |
| *Hospitalization for osteoporotic fractures/hip replacement surgery (negative control outcome)* | Log-binomial | -10.0 | (-33.1 to 8.6) |

Multivariable log-binomial regression models adjusted for age, sex, location of the health center in an area in needs of social transformation, institutionalization, baseline chronic diseases (heart failure, dementia, neurological/neuromuscular disorders and cerebrovascular disease), number of visits to primary care in the previous five years, and week of vaccination. CI: confidence interval. HD-IIV: high-dose, inactivated influenza vaccine. rVE: relative vaccine effectiveness. SD-IIV: standard-dose, inactivated influenza vaccines. *Poisson regression models were used when log-binomial regression models failed to converge.
